# Development of a Multi-Trait Polygenic Score for Intrinsic Capacity

**DOI:** 10.64898/2026.02.25.26347054

**Authors:** Melkamu Bedimo Beyene, Renuka Visvanathan, Robel Alemu, Nigussie T. Sharew, Olga Theou, Beben Benyamin, Matteo Cesari, John Beard, Azmeraw T. Amare

## Abstract

**Background:** Intrinsic capacity (IC) is a key marker of healthy ageing, which captures an individual’s physical and mental capacities, measured across five domains: cognitive, locomotor, psychological, vitality, and sensory. Although genetic factors are known to influence both general IC and its individual domains, existing IC indices have been developed primarily using phenotypic data, without accounting for the underlying biological architecture across domains. In this study, we developed a multi-trait polygenic score (Mt-PGS) model for IC by integrating polygenic scores derived from a broad set of phenotypes spanning the five IC domains and examined its validity.

**Methods:** Using data from 13,085 participants of the Canadian Longitudinal Study on Aging (CLSA), we computed PGSs for 63 phenotypes related to IC domains. A supervised machine-learning model was applied to develop a mt-PGS model for IC and identify the optimal set of polygenic predictors. The validity of the mt-PGS IC score was evaluated by comparing it with a phenotype-based IC score and by examining its association with mortality.

**Results:** Our analysis identified PGSs for 33 phenotypes with non-zero coefficients, jointly explaining 2.23% of the variance in IC. Several of the strongest contributors were most closely aligned with vitality-related phenotypes in the literature (including body mass index, grip strength, fat-free mass, diastolic blood pressure, and chronic obstructive pulmonary disease), acknowledging cross-domain relevance, and that predictors from all five IC domains were represented. The mt-PGS IC score was consistent with the phenotype-based IC score, positively correlated with the phenotype-based IC score and was inversely associated with mortality (OR = 0.04; 95% CI: 0.005 – 0.379).

**Conclusion:** Our findings support the multisystem biological basis of IC, demonstrating that an mt-PGS model integrating diverse phenotypes is associated with the phenotype-based IC score. PGSs for the phenotypes frequently related to vitality in the literature were the strongest predictors, recognizing that several of these phenotypes may span multiple domains, and that all domains contributed to the model. If replicated across different ancestries and settings, these findings may serve as a foundation for future research for the potential integration of genetic information into IC frameworks.

## Introduction

Intrinsic capacity (IC) represents the composite of an individual’s physical and mental capacities that underpin functional ability and contribute to ageing, as proposed by the World Health Organization (WHO) [1]. IC encompasses five domains: cognition, psychological well-being, locomotion, vitality, and sensory function that collectively reflect the body’s intrinsic reserve and resilience to age-related decline [2]. A growing body of evidence shows that IC predicts frailty [2-4], disability and functional dependence [3-7], and mortality [2, 4, 6, 7] more effectively than chronological age alone [6, 7].

Despite increasing consensus on the significance of IC as a comprehensive marker of healthy aging, its conceptual framework continues to evolve, and key questions remain to be answered. One of these questions is that current IC models rely on phenotypic information, derived from clinical assessments and/or self-reported measures that vary widely in content and degree of objectivity [8-12]. While these tools are valuable for capturing observable aspects of an individual’s functional ability, they do not consider the subtle physiological processes that precede perceivable or visible changes relevant to IC. Integrating genetic information into IC frameworks may, therefore, offer an opportunity to improve the precision of IC measurements and to strengthen the biological validity of this indicator.

The second key question concerns the relative contribution of each of the five domains to the overall IC, as well as whether a hierarchical structure exists in which one domain underpins the others. Whilst the broader literature shows that IC is a multidimensional construct with distinct contributions from five interrelated domains [3, 8, 9, 11, 13, 14], recent evidence suggests that these domains may not contribute equally to the general IC [15, 16]. In particular, the vitality domain has been proposed as foundational to IC, given its strong associations with core biological processes involved in ageing, including metabolic regulation, inflammatory pathways, and mitochondrial function [15, 17].

Our previous work, leveraging large-scale phenotypic and genetic data from the UK Biobank and the Canadian Longitudinal Study on Aging (CLSA), has demonstrated that IC is a complex, heritable trait [18]. In the first genome-wide association study (GWAS) of IC globally, 10 novel genomic loci and biological pathways associated with IC were identified [18]. Furthermore, lifestyle and socioeconomic factors, as well as their interactions with genetic susceptibility captured by polygenic scores (PGSs), were shown to influence IC, with an IC-specific PGS explaining a modest proportion of phenotypic variance [19]. However, because IC is a composite of multiple measures across the various domains, traditional single□PGS approaches, which focus on individual phenotypes, may overlook its multidimensional biological architecture. In this context, multi-polygenic score (mt-PGS) models, which integrate polygenic predictors from multiple genetically correlated traits, have emerged as a promising framework for characterizing complex phenotypes such as IC. Such approaches have consistently been reported to improve prediction performance across diverse outcomes [20-23]. For example, cardiometabolic risk prediction has been enhanced through the integration of lipid, metabolic, and inflammatory PGSs [24, 25], and the prediction of brain ageing phenotypes has been improved by combining cognitive and neuropsychiatric PGSs[26, 27].

In this study, an mt□PGS model was developed for IC in the CLSA by integrating PGSs from a broad set of phenotypes across the five domains, and its validity was evaluated by examining consistency with phenotype-based IC score and its association with mortality.

## Methods

### Study Cohort

CLSA is a large nationally representative, population-based cohort of adults aged 45–85 years at baseline (2010–2015), designed to examine biological, social, and environmental determinants of healthy ageing [28]. The CLSA included over 50,000 participants in total, subdivided into two subgroups [28, 29]: the tracking cohort (N∼20,000 participants) in which data were obtained through structured telephone interviews conducted across the ten provinces of Canada, and the comprehensive cohort (over 30,000 participants), in which data were collected through in-person physical assessment and the collection of blood and urine specimens at 11 designated data collection centres.

For this analysis, 13,085 participants from the comprehensive cohort, who had both genetic and phenotypic data, were included [18, 30].

### Target outcome: Intrinsic capacity score

The IC score was previously developed [18] using a factor analysis methodology applied to 14 baseline variables with high loading on the IC general factor. The validity of this IC score was examined by assessing its associations with age, sex and mortality.

### Development of a multi-trait polygenic score model for IC

The mt-PGS model for IC was developed using supervised machine learning methods in a stepwise manner (Figure 1) as outlined below.

**Figure 1:**
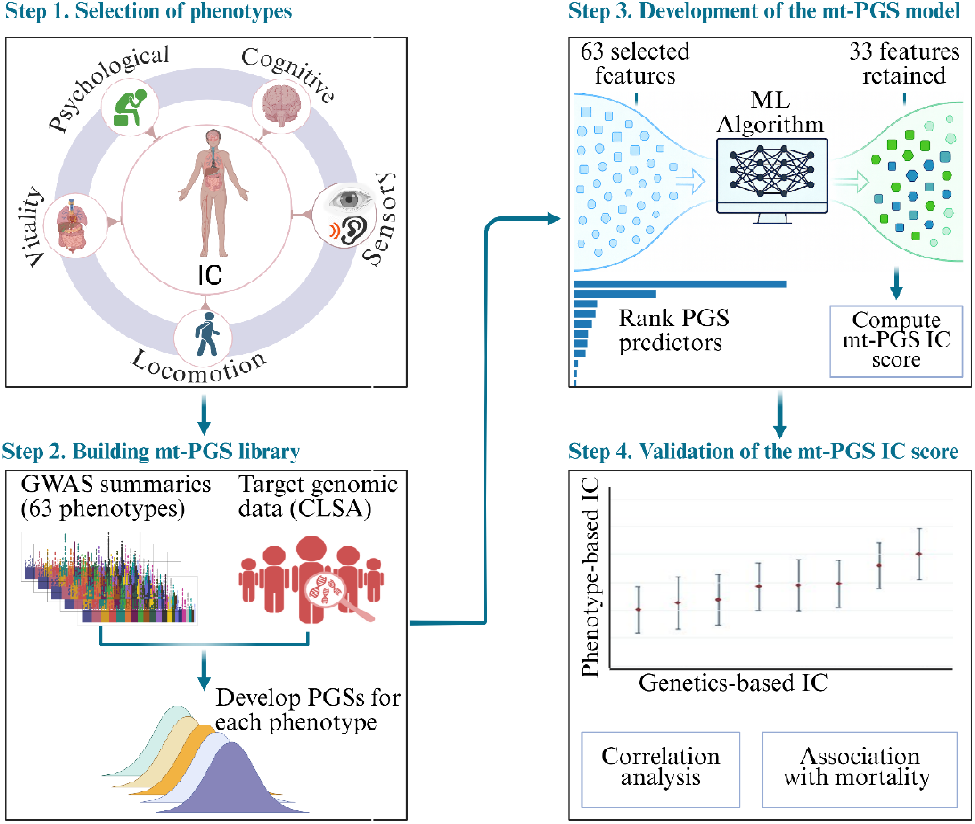
Schematic flow diagram of mt-PGS model development for IC. Step 1 involved the selection of phenotypes representing the five IC domains. In step 2, GWAS summary statistic were downloaded from public databases and quality control was performed. This was followed by the computation of PGSs (for each of the 63 selected phenotypes) in the target cohort and building a PGS library. In step 3, the mt-PGS model was fitted using machine learning methods, and mt-PGS scores were calculated. In step 4, we examined the validity of the mt-PGS IC score by assessing its correlation with the phenotype-based IC score, as well as its association with mortality. ***Abbreviations:*** *IC = Intrinsic capacity, GWAS = Genome-wide association study, ML = Machine learning, PGS = Polygenic scores, CLSA = Canadian longitudinal study on aging, mt-PGS = multi-trait polygenic scores*.

#### Step 1 – Selection of phenotypes

Based on our prior literature review [8] and empirical evidence on their association with IC [8, 18, 31], we identified 63 phenotypes likely related to the five IC domains. The GWAS summary statistics for these phenotypes (discovery sample) were then extracted (prioritizing recent studies with a larger sample size) from publicly available resources. The list of the 63 phenotypes, including the link to the full GWAS summaries, is provided in the Supplementary material.

To avoid sample overlaps and ensure independence between the discovery and target cohorts (an assumption of polygenic modeling), GWASs involving the CLSA cohort were excluded.

#### Step 2 – Building mt-PGS library

The PGSs for each phenotype were constructed using the Polygenic Risk Score-Continuous Shrinkage (PRS-CS) method [32], a Bayesian regression framework that infers posterior effect sizes from GWAS summary statistics while accounting for linkage disequilibrium (LD) across the genome. PRS-CS employs continuous shrinkage priors (CS) on SNP effect sizes, which enable adaptive shrinkage of estimated effects depending on the strength of the association signal and the local LD structure. For the current analysis, the GWAS summary statistics for each phenotype and the precomputed LD pattern of the 1000 Genomes European reference panel [33] were used to estimate posterior effect sizes and compute individual-level PGSs in the target CLSA cohort. All PGSs were standardized (transformed to a z-distribution) before the regression analyses.

#### Step 3: Development of the mt-PGS model

The mt-PGS model was developed in the CLSA using an elastic net supervised machine learning algorithm, implemented in R packages – the *glmnet* (for penalized regression), *caret* (for data preprocessing), *nestedcv* (for model validation and performance estimation), *and tidyverse* (for data management and visualization) packages in R (version 4.3.2) [34-37]. We used a five-fold nested cross-validation (nestedcv) framework to tune model hyperparameters and evaluate the predictive performance while avoiding data leakage between model training and testing [37, 38]. Within the inner cross-validation loop, we varied an elastic net mixing parameter (α) from 0.05 to 1.00 in increments of 0.05 to determine the optimal balance between ridge and LASSO penalties. For each α value, *glmnet* generated a sequence of regularisation parameters (λ) and the value that minimised the mean squared error (λ-min) within the inner folds was selected. All features were internally standardized (transformed to a z-distribution) within *glmnet* before model fitting. After training the model on 80% of the sample (inner loop), the final model was tested on the outer loop dataset (20% holdout test data), at the optimal α and λ values, and the model’s performance was evaluated using the coefficient of determination (R^2^), Root Mean Square Error (RMSE) and Mean Absolute Error (MAE). Finally, an elastic net regression model was refitted using PGSs with non□zero coefficients (33 features), and the mt□PGS IC score was generated in the 20% test sample.

The PGSs contributing most strongly to the mt□PGS were identified, and their relative contributions were characterized.

#### Step 4: Validation of the mt-PGS IC score

The genetics□based IC score (mt-PGS IC Z-score) developed in step 3 was validated by comparison with the previously established phenotype□based IC Z-score [18]. This involved correlation analysis between the two scores and assessment of the association between the mt□PGS IC Z-score and mortality. Associations with mortality were examined using a binary logistic regression model adjusted for age, sex, and the top five PCs.

## Results

### Sample characteristics

We analyzed data from a total of 13,085 participants, with a mean age (SD) of 61 (9.6) years. About half (50.8%) were female, and the majority (97.4%) had European genetic ancestry.

### Multi-trait polygenic model for IC

Of the 63 PGSs included as features in the machine□learning model, 33 had non-zero coefficients and therefore contributed to the mt-PGS IC model. The final mt-PGS model (Figure 2) explained 2.23% of the interindividual variability in IC (RMSE = 0.98 and MAE = 0.79).

**Figure 2.**
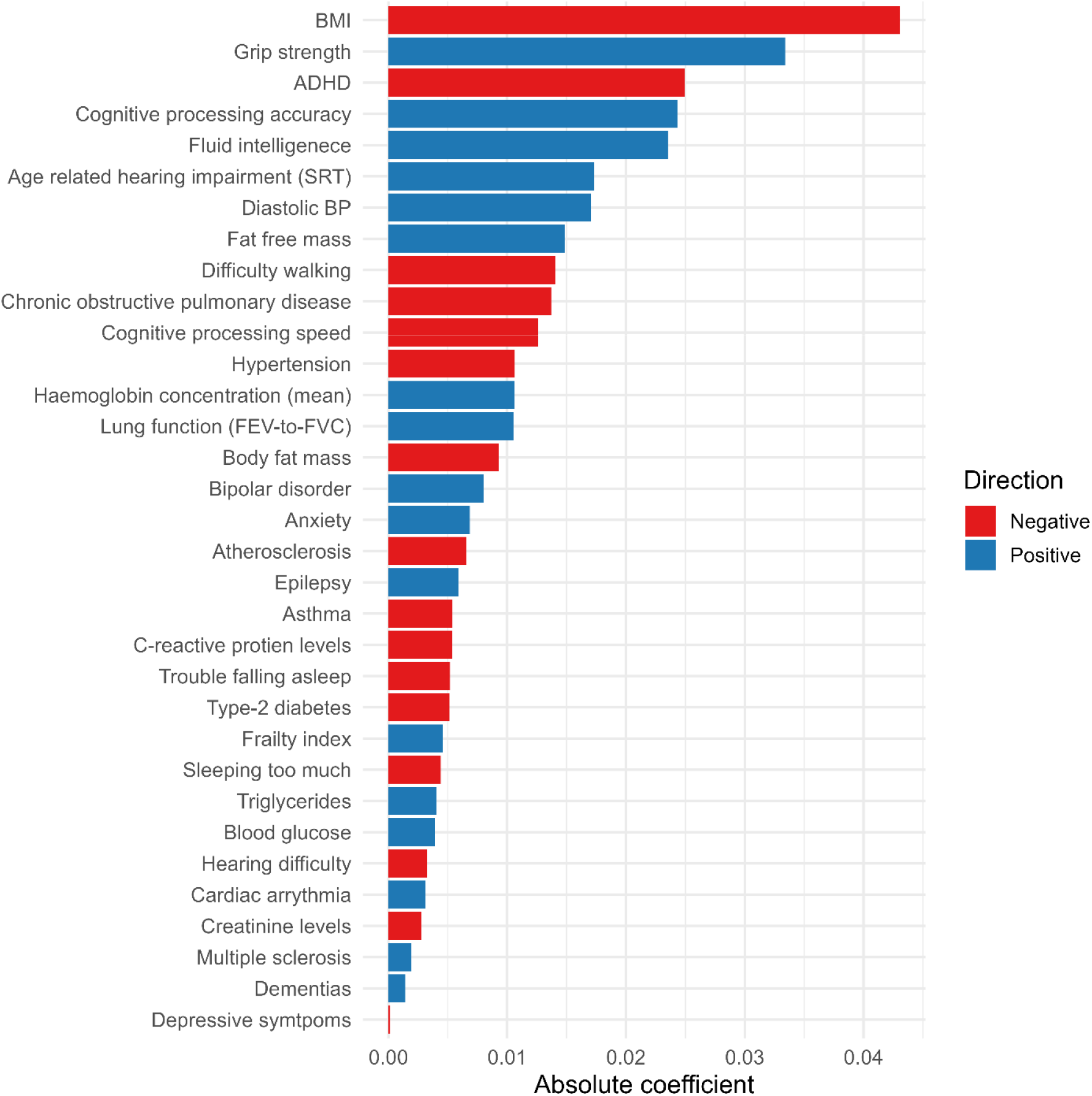
The 33 Features (PGSs) retained in the mt-PGS IC model. Bar lengths represent the absolute magnitude of standardized regression coefficients, and color indicates the direction of association (positive = blue, negative = red). ***Abbreviations:*** *BMI = body mass index, ADHD = Attention Deficit Hyperactivity Disorder, BP = blood pressure, FEV-to-FVC = ratio of forced expiratory volume to forced vital capacity*.

Based on their most frequent alignment in literature, while acknowledging cross domain overlap of the phenotypes, several of the strongest polygenic contributors to the mt-PGS IC model were closely aligned with vitality domain, including body mass index (BMI) (β = -0.04), grip strength (β = 0.03), diastolic blood pressure (β = 0.017), fat-free mass (β = 0.015), and chronic obstructive pulmonary disease (β = -0.014), in descending order of contribution. Importantly, there were also strong contributors retained from the psychological (ADHD: β = -0.025), cognitive (cognitive processing accuracy: β = 0.024 and fluid intelligence: β = 0.023), sensory (age-related hearing impairment: β = 0.017) and locomotion (difficulty walking: β = -0.014) domains (Figure 2). Full results are presented in Figure 2, with further details in the supplementary material.

### Validity of the mt-PGS IC score

Our results showed a consistent positive relationship between the mt□PGS IC score and the corresponding phenotype□based IC score (Figure 3). A modest but statistically significant positive correlation was observed between the two measures (r = 0.16; 95% CI: 0.12-0.19).

**Figure 3:**
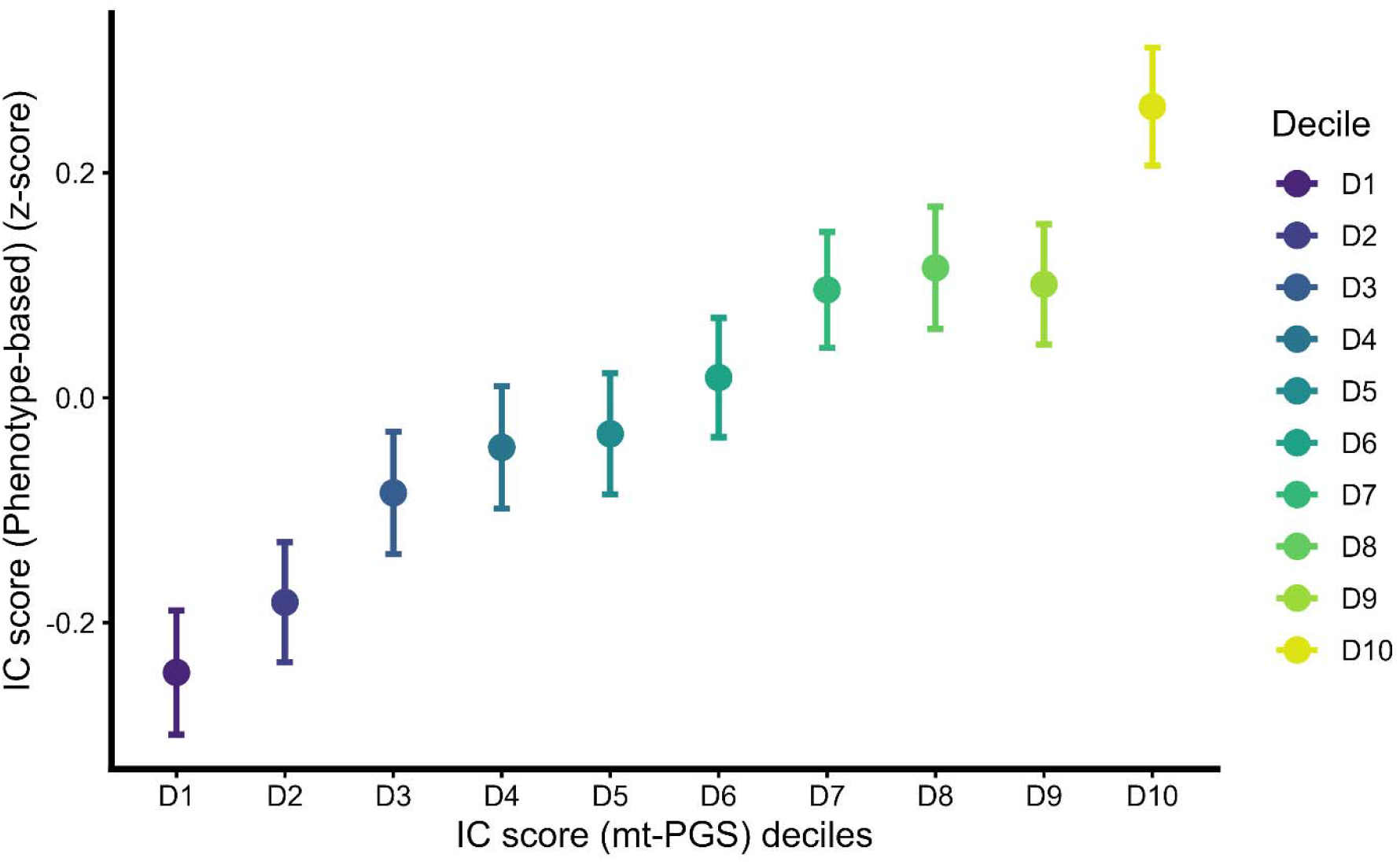
Relationship between phenotype-based IC score and mt-PGS IC score. Mean phenotype-based IC z-scores (y-axis) are shown across deciles of the mt-PGS-derived IC score (x-axis; D1–D10). Points represent the mean phenotype-derived IC Z-score values at each decile of mt-PGS values, and vertical error bars indicate 95% confidence intervals. Colors correspond to mt-PGS deciles. ***Abbreviations***: *IC = Intrinsic capacity, mt-PGS = multi-trait polygenic score*.

Risk (proportion) of death also decreased with increasing tertiles of both phenotype-based and genetically derived IC scores, as shown in the supplementary material. Logistic regression analysis result also confirmed this inverse association, that higher mt-PGS IC Z-scores were associated with lower odds of death (adjusted OR = 0.04; 95% CI: 0.005 – 0.379), corresponding to an approximately 96% reduction in mortality risk per one Z-score increase.

## Discussion

This is the first study to develop a multi-trait polygenic score (mt-PGS) model for IC, providing new insights into a genetically informed IC measure. Genetic scores representing all five domains were retained among the top ten contributors. The two strongest predictors were most frequently aligned with vitality-related phenotypes in the literature, although these phenotypes likely reflect overlapping biological processes across domains. The mt-PGS IC score was positively correlated with phenotype-based IC and inversely associated with mortality. These findings are consistent with the conceptual framework of IC, reflecting the body’s intrinsic reserve across multiple biological systems and that aggregating polygenic information from multiple phenotypes results in a more informative genetic index than single-PGS approaches.

The use of mt-PGSs in this study increased the variance explained in IC by more than 7-fold (to 2.23%), from 0.3% using a single PGS (i.e. IC-PGS) in our previous study [19]. This enhanced predictive performance is in line with findings related to mt-PGS studies for other health conditions [21, 23, 39, 40], with great improvement in predictive performance of up to 9-fold in some psychiatric conditions, such as ADHD [23, 41]. Although the variance explained by the mt-PGS is modest when considered in isolation, it is comparable to other multi-trait polygenic prediction models for complex ageing-related traits. Such genetic information may have potential value when integrated with phenotypic determinants, although further work is required to quantify its incremental predictive contribution over established assessments. A growing body of evidence demonstrates that integrating polygenic risk scores with phenotypic or clinical risk factors improves risk stratification and decision-making across a range of health outcomes. For example, the addition of PGSs to established clinical risk models [42] has been shown to refine cardiovascular risk prediction beyond traditional factors alone [43, 44], improve identification of individuals at high risk of type 2 diabetes when combined with demographic and metabolic indicators [45], and enhance breast cancer risk stratification when incorporated into comprehensive models alongside family history and clinical variables [46, 47]. With this context, integrating genetic information into IC assessment may complement the phenotype-based IC measures by capturing underlying biological vulnerability, thereby supporting earlier identification, monitoring, and assessment of functional decline and enabling timely preventive or targeted interventions before overt impairment becomes evident.

One key finding from this study was that several of the strongest polygenic contributors, such as BMI, grip strength, fat-free mass, diastolic blood pressure, and chronic obstructive pulmonary disease (COPD), have often been linked to vitality in prior literature, although they may relate to other domains. This pattern may likely point towards the central role of this domain and related physiological reserve and metabolic function as core components of IC [48]. These findings align with evidence documenting that genetic determinants of BMI and body composition influence inflammatory regulation, energy metabolism, and hormonal pathways that modulate resilience and healthy ageing [49-51]. Grip strength, mostly used as a marker of overall vitality, but also for locomotion, has been shown to have high heritability and strong predictive power for mortality, frailty, and disability [52-54]. Similarly, fat-free mass reflects muscular and metabolic integrity, closely related to both vitality and locomotion function, supporting the body’s capacity to maintain homeostasis and functional reserve [55, 56]. The contribution of blood pressure and COPD to the model is in line with the WHO’s working definition of vitality [15] and may further emphasize the role of cardiovascular and pulmonary physiology in supporting IC, consistent with evidence that reduced vascular and respiratory physiological functions accelerate biological ageing and limit activity tolerance [57, 58]. In addition to vitality, function-related PGSs, such as cognitive processing accuracy and fluid intelligence and locomotion-related phenotypes, such as difficulty walking, in addition to grip, were also among the strong contributors, with the other two domains represented in the top ten as well, reinforcing the multifactorial nature and multisystem genetic architecture of IC [3, 6, 31], though they may have disproportionate contributions. Collectively, predictors most often aligned with vitality were prominent among the top contributors, while cognitive and locomotor domains were also strongly represented, and other domains contributed. This pattern is consistent with a potentially prominent role of vitality-related processes within IC [16], although further work is required to confirm this.

For most phenotypes, the direction of association between the PGSs and IC was consistent with that observed at the phenotypic level, although this pattern was not uniform across all traits. For example, while the phenotype-based IC was lower among individuals with bipolar disorder, the corresponding PGS showed a small positive association. A possible explanation for this discrepancy is that the phenotype-based IC reflects the direct consequence of the disorder, while the PGS captures biological liability that may also be associated with other phenotypes positively related to IC. Consistent with this interpretation, prior studies have demonstrated that bipolar disorder genetic liability is positively associated with higher physical activity levels [59] and greater creativity [59] and cognitive performance [60], and is genetically correlated with higher socioeconomic status [61], all of which are positively related to IC. In general, similar mechanisms may underlie other traits for which associations between PGSs and IC contrast with those observed at the phenotypic level.

The consistent pattern of relationship between the mt-PGS-derived IC score and the already established phenotype-based scores confirms the validity of the genetically informed IC measure as an index for IC. This is further supported by the inverse association of the mt-PGS-based IC score with mortality, characterized by a higher risk of death among those with a lower mt-PGS-based IC score – 96% lower risk of death for a one Z-score increase in mt-PGS IC score. A similar pattern has been reported for phenotype-based IC, with extensive evidence, including our previous study, demonstrating that lower phenotype-based IC is associated with increased mortality risk [6-8, 18, 31].

While this study provided interesting insights into the genetically informed IC score, some consideration should be taken when interpreting the findings. First, the study used samples of a predominantly European ancestry population, which may limit transferability to other ancestries. Second, the 63 inputs varied in GWAS sample size and quality, which may have introduced differential weighting to phenotypes with larger-sized discovery datasets. Third, our target cohort is modest for genetic analyses that often requires larger sample to optimally estimate effect sizes. Fourth, although nested cross-validation minimized overfitting, external validation in independent and ethnically diverse cohorts is essential to confirm the generalizability and robustness of these findings. Finally, it should be acknowledged that the classification of phenotypes into IC domains was guided by existing literature and does not imply strict exclusivity; several phenotypes likely reflect overlapping biological processes and may contribute to more than one domain.

In summary, this study provides the first genetically informed index for IC that integrates PGS across multiple IC-related phenotypes and demonstrates its consistency with the phenotype-based IC measure and association with mortality. Combining polygenic predictors across various body systems spanning the domains of IC could support future research for the possible integration of genetic susceptibility into the IC framework, although further work is required to validate predictive performance and establish its clinical utility. Our findings serve as a foundation for further research into whether integrating IC-related mt-PGSs with lifestyle, environmental, and multi-omics factors improves the explanatory or predictive modelling of IC.

## Acknowledgments

This research was made possible using the data/biospecimens collected by the CLSA. Funding for the CLSA is provided by the Government of Canada through the Canadian Institutes of Health Research (CIHR) under grant reference: LSA 94473 and the Canada Foundation for Innovation, as well as the following provinces, Newfoundland, Nova Scotia, Quebec, Ontario, Manitoba, Alberta, and British Columbia. This research has been conducted using the CLSA dataset, Comprehensive baseline v7.0 and GEN 3 data sets under Application Number [2304009]. The CLSA is led by Drs. Parminder Raina, Christina Wolfson and Susan Kirkland. The Authors gratefully acknowledge the time and commitment of the CLSA participants, without whom this research would not be possible.

## Declarations

Professor Renuka Visvanathan and Professor John R. Beard are members of the World Health Organisation Clinical Consortium of Healthy Ageing.

The authors declare that the authors alone are responsible for the views expressed in this article and do not necessarily represent the views, decisions, or policies of the institutions with which they are affiliated, nor the views of the CLSA.

## Data Availability

Data are available from the Canadian Longitudinal Study on Aging (www.clsa-elcv.ca) for researchers who meet the criteria for access to de-identified CLSA data, and the GWAS summary statistics are publicly available to anyone in the corresponding consortia from which we obtained them.

